# Integrative Machine Learning Approach to Risk Prediction for Dementia and Alzheimer’s Disease

**DOI:** 10.1101/2025.06.02.25328770

**Authors:** Amos Stern, Michal Linial

**Affiliations:** The Rachel and Selim Benin School of Computer Science and Engineering, The Hebrew University of Jerusalem, Jerusalem, Israel; Department of Biological Chemistry, The Life Science Institute, The Hebrew University of Jerusalem, Israel

**Keywords:** APOE, AUC, Feature selection, GWAS, PWAS, SHAP values, UK Biobank

## Abstract

**Background:** Dementia, especially Alzheimer’s disease (AD), is a major global health challenge marked by progressive cognitive impairment, behavioral changes, and loss of autonomy. As global life expectancy rises, there is a growing urgency for earlier diagnosis and better clinical guidance for AD and other dementia subtypes.

**Objectives:** This study aimed to develop and evaluate machine learning (ML) models for predicting AD risk using comprehensive health, genetic, and lifestyle data. It further sought to examine sex-specific model performance and the predictive potential for vascular dementia (VaD) versus non-vascular dementia.

**Methods:** Data from the UK Biobank (UKB) cohort were analyzed, comprising 2,878 individuals diagnosed with AD and 72,366 age-matched controls. Multiple ML algorithms were evaluated, with CatBoost achieving the highest performance (ROC-AUC = 0.773). Input features included ICD-10 medical codes reported five years prior to diagnosis, lifestyle and environmental factors, and genetic variants including the ApoE-ε4 alleles.

**Results:** CatBoost outperformed other ML approaches, with stronger predictive accuracy in females. VaD prediction was more accurate despite the smaller sample size. ApoE-ε4 was confirmed as a key genetic risk factor, while other genetic markers had limited predictive value. Significant non-genetic predictors included comorbid conditions (e.g., type 2 diabetes, hypertension), educational attainment, physical activity, diet, and cardiovascular health, highlighting the multifactorial nature of dementia risk.

**Conclusions:** The integration of genetic, clinical, and lifestyle data enhances the accuracy of AD and dementia risk prediction. Findings underscore the importance of sex-specific analysis and the influence of comorbidities and modifiable risk factors. This approach supports more precise, personalized early interventions and diagnostic strategies for AD and dementia subtypes.

**Highlights:**

◦ CatBoost accurately predicted Alzheimer’s disease risk, with higher performance in females.
◦ The ApoE-ε4 gene was the strongest genetic risk factor across AD and all dementia subtypes.
◦ Comorbidities (e.g., heart disease, diabetes) interact with lifestyle factors and education to increase the risk predictive models.
◦ Vascular dementia showed strong predictive signals despite smaller sample size.

## 1. Introduction

The prevalence of dementia, including Alzheimer’s disease (AD), is rapidly increasing due to global population aging [1, 2]. AD accounts for 60-70% of all dementia cases and is a leading cause of dementia in individuals aged 65 and older. Dementia is a global health concern, affecting over 55 million people, with nearly 10 million new cases annually. According to the World Health Organization (WHO), by 2050, the number of people living with dementia is projected to triple, reaching over 150 million globally [3]. Dementia significantly contributes to disability and dependency in older populations [4]. It is also among the leading causes of death worldwide and imposes an enormous burden on the medical system, as well as on health and caregiving economies. Key epidemiological trends show that the fastest growth in the occurrence of dementia is in low- and middle-income regions, which account for about two-thirds of dementia cases [5, 6]. Overall, the increase in prevalence and mortality rates associated with AD likely also reflects underdiagnosis, which applies across North America, Europe, and Asia [7]. Still, in some high-income countries, the increase in AD has been attenuated, attributed to improved education and healthcare, and to some extent, also to lifestyle changes [8]. Many factors, such as age and genetic predisposition, have been reported to increase the risk of developing AD [9, 10]. The Dementia prevention, intervention, and care committee (2020) identified 12 risk factors, including low education, hypertension, hearing loss, smoking, obesity, depression, physical inactivity, diabetes, and low social contact [11]. These factors account for about 40% of all cases. With the added roles of alcohol consumption, traumatic brain injury (TBI), air pollution, untreated vision loss, and high LDL cholesterol, the contribution of modifiable factors is even greater [12, 13].

AD is best described as a multiplex disorder involving multiple pathological pathways, including neuroinflammation [14], endocytosis, cholesterol transport, APP and tau processing, protein folding, and ubiquitination [15]. These processes lead to progressive, irreversible changes in brain structure and function, particularly affecting vascular systems and neuronal connectivity. This disrupts the interplay among neurons, microglia, astrocytes, and oligodendrocytes [16]. Clinically, AD features a prolonged preclinical phase during which anatomical and molecular alterations—such as β-amyloid plaques, immunoreactive senile plaques, and tau neurofibrillary tangles—develop silently [17, 18], long before cognitive symptoms like memory loss or disorientation emerge.

Genetically, early-onset familial AD (EOAD) involves pathogenic variants in APP, PSEN1, and PSEN2 [19, 20]. In late-onset cases, APOE variants, especially APOE4 (R112, R158) are key modulators of risk and pathology [21]. Genome-wide association studies (GWAS) have identified over 50 susceptibility loci, reinforcing the disease’s polygenic nature and the central role of APOE [22]. Mixed dementias, such as AD coexisting with vascular dementia (VaD), become more prevalent with age [23], with cerebral amyloid angiopathy (CAA) as a major contributor to vascular-related pathology [24]. Other dementias, including Lewy body disease and TDP-43 encephalopathy (LATE), complicate differential diagnosis and care.

Machine learning (ML) has enabled early prediction of AD using large-scale clinical datasets. Models integrating electronic health records (EHRs) and genetic profiles can forecast disease onset years in advance [25–27]. Enhancements using polygenic risk scores (PRS) [28], natural language processing (NLP) [29], and unsupervised deep learning [30] have improved performance in detecting mild cognitive impairment and predicting progression to dementia [31, 32].

Based on the elevated risk of dementia with age and the difficulty in defining the type of dementia and its severity, having a population-based, generalizable risk assessment protocol is crucial for the healthcare system and medical professionals. In this study, we integrate clinical data, leading genetic variants, and lifestyle information to estimate the relative importance of risk factors in predicting dementia, particularly Alzheimer’s disease. As age is clearly one of the most influential risk factors in all AD models, we neutralize its contribution, allowing us to reveal more subtle factors. We also provide alternative models for females and males and analyze the contribution of a wide variety of known and novel risk factors to improve diagnosis. This study highlights the understudied importance of modifiable risk factors, life experiences, and the relatively limited contribution of genetics beyond APOE to the risk of dementia and its various subtypes.

## 2. Materials and Methods

### 2.1. UK biobank (UKB) database, extraction and processing

The UKB includes over 500,000 participants collected from 23 medical centers across the UK, who were recruited over the years 2006–2010 for participants aged 40–69 (54.4% are females) [33]. All analyses were based on the 2021 UKB release. Disease classification is based on clinical information encoded by ICD-10 codes F00 (dementia in Alzheimer’s disease; AD) and G30 (AD). For AD, we expanded our definition to broader phenotypes which are defined as dementia (including VaD and nonvascular forms of dementia. vascular dementia is code F01, and we included all other diagnose codes for dementia: F00, F02 and F03).

To avoid biases due to overrepresentation of genetic factors, we removed genetic relatives by keeping only one representative of each kinship group. This resulted in a dataset that includes 469,637 participants. To mitigate the age factor, which is the strongest factor in onset of AD, a protocol for age-dependent matching of the AD and control groups was performed implying a stochastic matching process between the two groups. The objective is to keep the majority of the samples while matching the age of recruitment distribution. In practice, we randomly chose samples from the control group (a total of 72,366 individuals) with the same age distribution as the age distribution of samples with Alzheimer’s diagnosis (AD group, 2,878 individuals). The rest of the analysis was performed on the matched sets. We performed 10 iterations for each model training with different random seeds to generate different random controls.

### 2.2. Machine learning (ML) methodology

#### 2.2.1. Algorithm selection

We used a state-of-the-art machine learning (ML) gradient boosting tree model of CatBoost (Categorical Boosting) [34, 35]. CatBoost is a gradient boosting algorithm that is specifically optimized for datasets with categorical features. It handles categorical variables natively without requiring extensive preprocessing like one-hot encoding, and it employs ordered boosting and efficient use of permutations to reduce overfitting and prediction shift bias. In each step of the algorithm, a decision tree base learner is created, using the previous iterations’ decision tree residuals as a gradient for minimizing the current tree’s loss function.

#### 2.2.2. Data partitioning and cross-validation strategy

The age-matched dataset (n=75,244) was partitioned using stratified random sampling to maintain class balance across splits: training set (68%, n=51,157), validation set (12%, n=9,028), and test set (20%, n=15,047). To assess model stability and robustness, we performed 10 independent training iterations, each using different random seeds (seeds 0-9) that generated unique data partitions while maintaining the same split proportions. This approach allowed us to evaluate both model performance variability and feature importance consistency across different data configurations. When training the model, the majority of other parameters remained at CatBoost default values, which have demonstrated superior performance in comparative studies and reduce the risk of overfitting through automated parameter selection [36]. For hyperparameter configuration see Supplemental **Text S1**.

#### 2.2.3. Model training process

Training followed CatBoost’s ordered boosting approach, where each iteration builds a decision tree using gradient information from previous iterations’ residuals. For categorical features, CatBoost computes target statistics using ordered boosting with random permutations of the training data, effectively mapping categorical variables to continuous space while avoiding prediction shift. The validation set was used exclusively for early stopping decisions and hyperparameter evaluation, while the test set remained completely held out until final model assessment. As in UKB, the number of AD cases is <4% of the dataset, we addressed class imbalance by weighting the positive class by the ratio of negative to positive samples. To prevent overfitting, we used CatBoost’s early stopping feature [36] based on a separate validation set. Training stopped when no improvement was observed for a set number of iterations after the best metric value. Logistic regression models were also trained for comparison.

#### 2.2.4. Feature set variants

To investigate the relative contribution of different feature types, we trained multiple model variants labelled as follows: (i) <u>All features</u>: complete feature set including clinical, genetic, demographic, and lifestyle variables. This includes the 45 expert-based features, combined with genetics and diagnosis). (ii) <u>Selected</u> <u>ICD-10</u>: statistically significant ICD-10 codes (n=66) combined with genetic and demographic features. (iii) <u>Reduced:</u> minimal feature set including sex, education, genetic variants, and ICD-10 diagnoses. (iv) <u>Genetics</u> <u>only:</u> 10 selected SNPs based on GWAS associations that were detected in 100% of the participants. (v) <u>ICD-</u> <u>10 only:</u> clinical diagnosis codes without genetic or demographic information. (vi) <u>PWAS variants:</u> gene-based functional scores for top-ranked AD-associated genes. Each model variant was trained using identical data partitions and hyperparameter settings to ensure fair comparison of predictive performance across feature combinations. We trained additional models only on a subset of features (e.g., genetics, ICD-10 diagnoses, sex, and education). Moreover, we trained models on UKB population partitioned by (i) sex for male and female groups, (ii) education preprocessed to poor, high school, and academic degree, (iii) APOE variants with partition to the APOE_e4_e4 (homozygotes with e4-e4 alleles), no_APOE_e4_e4 (samples without e4-e4 alleles), APOE_e4_hetro (heterozygotes with a single e4 allele), and no_APOE_e4_hetro (samples without the e4 allele).

#### 2.2.5. Feature selection and engineering

We have included all listed ICD-10 diagnoses as features. we filtered out the ICD-10 diagnoses to retain only diagnoses that are dated back ≥ 5 years prior to the AD diagnosis, and ≥ 5 years before the mean diagnosis age of the relevant affected age-group for the control group. We created a selected ICD-10 diagnosis features model by selecting the top ICD-10 diagnosis by performing Fisher’s exact test based on the contingency tables on the training set of affected vs. control groups for each ICD-10 diagnosis term. We selected only the significant ICD-10 codes (FDR < 0.05) and were able to reduce the list to only 66 significant ICD-10 features. In addition to the data fields from the UKB, we engineered features which were not explicitly defined by the UKB (e.g., the mean of systolic and diastolic blood pressure calculated from the list of blood pressure samples). Extracted education values were simplified to 3 categories of poor, high school, and academic degree.

### 2.3. Genetics analysis

#### 2.3.1. Variant genetic analysis

The UKB genotyping scheme is based on 805,426 preselected genetic variations which are expended by an imputation protocol to approximately 9M variants that passed quality control [37]. We used the Open Targets Genetics (OTG) platform [38] to select the most updated genetic associations for AD. OTG compiled the top-scored variants summary statistics from the GWAS catalog [39]. For top listed candidate genes from OTG we included the functional damaged score based on FIRM [40], as implemented by PWAS [41, 42]. We generated a list of 97 unfiltered SNPs and chose the 10 most frequent SNPs from the UKB dataset. The gene scoring methods by PWAS is described in Supplementary **Text S3.**

In addition, we used the in-house PRS and the UKB provided PRS [43]. We used LDpred2 to derive the AD PRS from UKB. In addition, we used the previously published PRS (v.2) from UKB based on the GWAS summary statistics and the imputed genotype data [44, 45]). Note that the optimal practice for AD PRS is sensitive to the SNP selection thresholds [46].

### 2.4. Dementia models and subtypes

One of the goals was to assess the variation in the risk factors according to different clinical terms associated with dementia. Several models were trained on the different individuals with dementia by their labels. For dementia group we used the ICD-10 diagnosis of F00, F01, F02 and F03 as positive. For the vascular dementia, samples with ICD-10 F01 were labeled as positive. For the unique vascular dementia, samples with the F01 ICD-10 but not any of the other dementia diagnosis (F00, F02 or F03) are labeled as positive. For the non-vascular dementia, samples that have a dementia ICD-10 diagnosis that is not vascular dementia (e.g., samples with F00 or F02 or F03 and no occurrence of F01.

### 2.5. Model evaluation and statistics

Model performance was primarily assessed using the area under the receiver operating characteristic curve (ROC-AUC), which measures the model’s ability to distinguish between classes across thresholds. We used SHAP (SHapley Additive exPlanations) to estimate the features’ importance [47]. By assigning a consistent importance value to each variable, SHAP results are presented as a rank list of the most strongly influence features with respect to the original values of each feature. We estimated the model performance also by other metrics including accuracy, precision, recall, and F1 score.

In comparing two groups (e.g., case and controls), and testing for statistically significant difference in the variable, we applied both t-test and the Mann-Whitney U test. To test the altered distributions between groups, we applied Kolmogorov-Smirnov (KS) test that is used to estimate whether the two samples come from the same distribution. We applied the Fisher’s exact test that yields exact p-value to determine whether there is a non-random association between two categorical variables (i.e., 2x2 contingency table).

## 3. Results

### 3.1. Balancing the age effect from AD group by a matching protocol

The primary goal of this study was to review current risk factor knowledge and evaluate its contribution to AD and dementia prediction in general. As a population-based resource, the UKB is based on standardized data collection protocols (see Materials and Methods). The average age of the patients in UKB is 57.1 years old (standard deviation, Std 7.66). We have retrospectively analyzed personalized clinical information on diagnosis, medical procedures, lifestyle, personal genetics, self-reporting, and nurse interview reports. As the strongest risk factor for AD is age, we have activated an age matching protocol to remove the impact of this feature by imposing an age-match of cases diagnosed with AD and control groups.

**Fig. 1A** shows the distribution of the participants in the study for controls and AD diagnosed (AD group). The Kolmogorov-Smirnov (KS) test confirmed the significant difference in the age distribution among AD group and controls (p-value <e-300). Repeating the statistical tests following the matching protocol resulted in an insignificant difference between the two groups (KS, p-value 1.0; **Fig. 1B**). Additional statistical tests including t-test and Mann-Whitney U test confirmed that the age feature was cancelled out for the AD (Supplementary **Table S1**). The rest of the analysis was performed on the age-matched data. Note that such matching protocol (See Supplemental **Text S2**) is used to avoid the many factors that are implicitly age-dependent, including the occurrences of age-dependent comorbidities.

**Figure 1.**
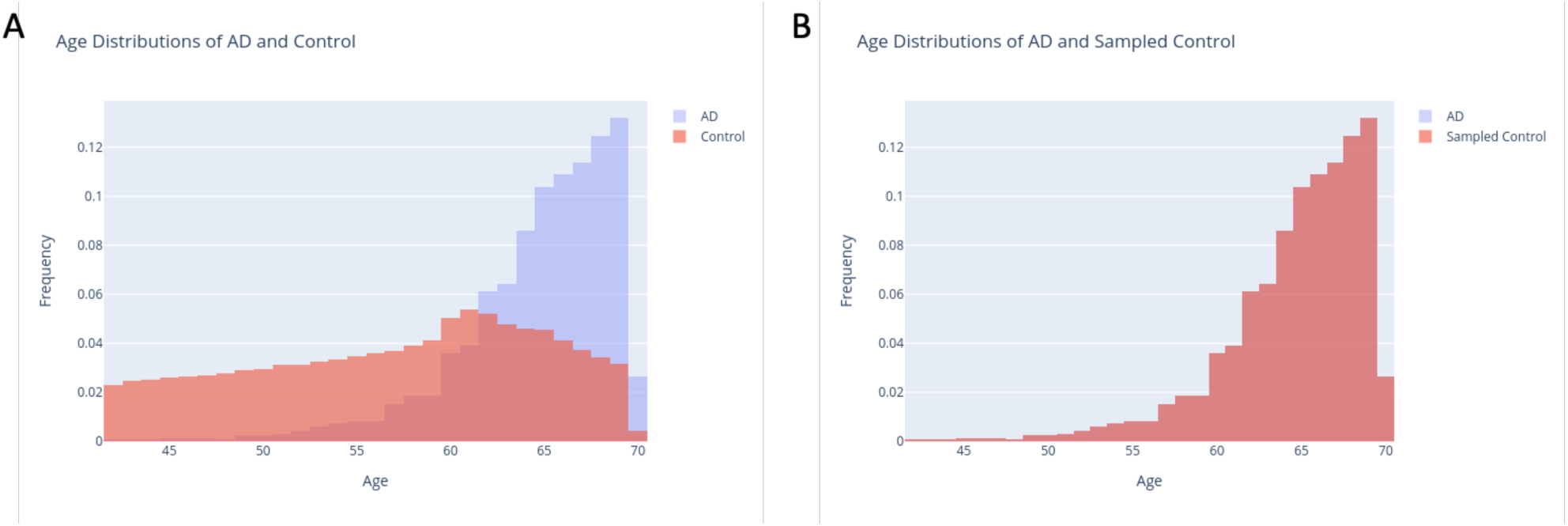
Age matching protocol. **(A)** The distribution of the control and AD groups by age. **(B)** Following a protocol for age-matching schemes, a major cofounding bias was removed, and each age a matched proportion of control and AD-groups remains stable throughout. For the matching algorithm, see Supplementary **Text S2**.

### 3.2. Clinical view using a time stamp for AD comorbidity

We performed Fisher’s exact test on the ICD-10 diagnosis of AD vs. the control groups. ICD-10 diagnoses were filtered to only include diagnosis 5 years prior to AD diagnosis for the affected individuals and 5 years prior for the mean age of the diagnosis for the control groups (see Materials and Methods). The relatively extended time-stamp (5-years) is used to make sure that administrative delay in AD diagnosis is likely to be minimal and therefore, we can properly separate comorbidities that tend to increase with age. Top diagnoses were used as features for the “selected icd10” models.

Supplementary **Fig. S1** shows that while most ICD-10 are insignificant and do not meet the statistically significant of FDR ≤0.05, a few significant protective ICD-10 codes are shown on the negative side of the odds ratio (OR <0) axis. In contrast, many more ICD-10 diagnoses tend to increase AD risk (the graph is shifted to the positive values of log2(OR). The most significant ICD-10 that overlap with AD are listed in Supplementary **Table S2**. Notably, the ICD-10 I25.9 diagnosis of a chronic ischemic heart disease (unspecified) shows an OR of 3.47 and a very strong z-value (1.23).

### 3.3. AD and dementia model performance

The relatively limited size of the AD group (filtered to avoid duplications based on multiple dementia annotations, insufficient support for variant calling etc.) encouraged us to carefully select a controlled number of features for each model. To minimize the number of selected features, we restricted the analysis to 45 features called *“all”* including BMI, smoking habits, cholesterol levels, blood pressure, playing computer, and other features that were used in consulting with the CNP lab in Hadassah Medical Center (Jerusalem, Israel). In addition, we included 10 selected SNPs and all ICD-10 diagnoses as one text feature. *“Selected ICD10”* is a collection of top 66 ICD-10 combined with genetic features. *“Reduced”* refers to the sex and education with the genetic features and ICD-10 diagnoses. *“Only genetic”* includes the 10 selected SNPs that are specified as the common variants with the highest coverage among all individuals in UKB. *“Only selected”* are the 66 significant ICD-10 that were selected as features. *“PWAS-global”* includes the results from PWAS-based scores (total 40, that covers inheritance modes of recessive and dominant) for the top 20 genes AD gene associations from the OT global score. *“PWAS-genetics”* includes the 18 genes from the top OTG score according to PWAS scores (total 36, including the recessive and dominant modes). We performed the model training for the different population subsets (see Materials and Methods). The summary of the models’ performance metrics including the robustness assessment of each model (i.e., Std) can be viewed in **Fig. 2A**.

**Figure 2.**
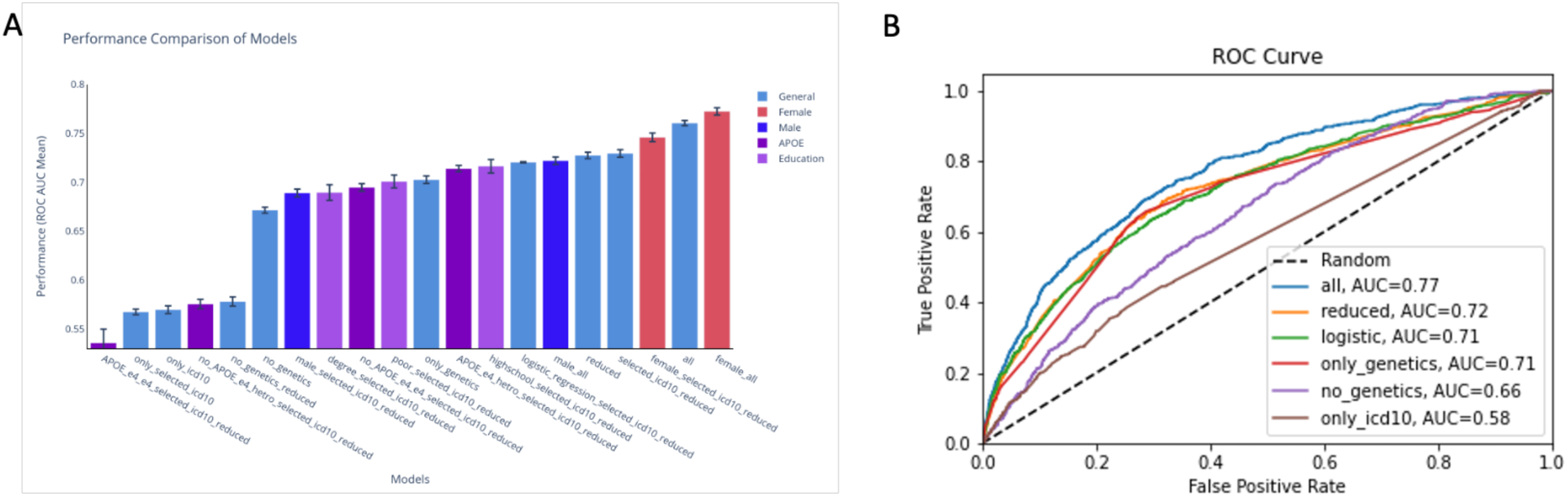
Performance of the risk factor predictive modes for AD from UKB**. (A)** Comparison of selected models’ performance by the mean of the ROC-AUC for 10 different independent training iterations. Error bars present the standard errors of the model’s replication. Bars are colored by their category: general, female, male, APOE and education. **(B)** Comparison of the performance of selected AD prediction models. The diagonal line marks a random no discrimination line (AUC 0.5). Detailed on the performance and statistics of related models that included the OT gene list and the PWAS scoring is available in Supplementary **Table S3**.

The model with the best ROC-AUC score metric is for *“female_all”* which is even higher than the one that includes all features for the unified samples from both sexes. The male models ROC-AUC scores are significantly lower, suggesting different manifestations of AD in males and females. Visually, one can see that there are 2 main classes of performances, the lower performance group (ROC-AUC ∼0.6) suffer from relatively small group size, and consequently a low statistical power (e.g., *“APOE_e4_e4_selected_icd10_reduced”*). Models that included only ICD-10 codes were limited to genetic features *(“only_icd10”, “only_genetics”*) and resulted in lower performance with ROC-AUC of ∼0.55 and 0.58, respectively. We conclude that while genetic and ICD-10 data contribute to model performance, they may not be sufficient without the contribution of rich demographic and clinical features. The other group of models is substantially better (ROC-AUC of ≥0.67). The population sizes and the performance metrics including precision, recall and accuracy are available in Supplementary **Table S3.**

Considering only genetic features hinders the model ROC-AUC to ∼0.7, while removing any genetic-origin features reduced the performance to ROC-AUC of ∼0.67. The reduced models are less predictive than the *“all”* models across all of the different groups. For the *“reduced”* model using logistic regression (LR) instead of CatBoost reduced the performance, emphasize the contribution of non-linear relations and feature interactions (**Fig. 2B**). Using the 66 selected ICD-10 codes of features instead of all of the ICD-10 data, does not affect the performance substantially, the “*all”* and “*selected_icd10_all models”* ROC_AUC was quite stable. Using the reduced features instead of the expert-based features led to a drop in ROC-AUC, a drop from 0.761 *(“all”)* to 0.728 (*“all_reduced”).* The drop in ROC-AUC score is consistent across all model groups (see Supplementary **Table S3**).

Interestingly, for the education models the high-school models showed better performance than the poor and academic degree models. This can be explained by the size of the positive labelled samples from the total (1,227 vs 554). To further test the stability of the results, we performed down-sampling and bootstrapping to neutralize the direct effect of the group size, and to test robustness. Based on such manipulation in the data size, we conclude that there is no statistically significant difference between the means of the educational partitioned groups.

### 3.4. Gene-based PWAS score failed to boost AD risk predictive models

As seen in the ROC-AUC curves (**Fig. 2B**), the *“all”* model outperforms the other selected models in all ranges while *“only_genetics”* and *“only_ICD10”* performed relatively poorly. We conclude that interaction of genetics, clinical and other features are critical for AD risk prediction. For the ICD-10 diagnosis features, we applied a cutoff of 5 years prior to the age of diagnosis. Interestingly, repeating the same protocol but reducing the cutoff to only one year prior to diagnosis age, the model metrics improved significantly. For example, for the *“all”* features model the ROC-AUC improved from 0.761 (5 years cutoff) to 0.81 (1 year cutoff).

We tested the possible contribution PWAS as a gene-based association method [48]. PWAS is a complementary approach for routine GWAS based on FIRM scores that assess the effect of genetic variants on gene function per individual (see Supplementary **Text S3**). We selected two groups of genes as features for the models training. The first model group called *“PWAS-global”* was composed of the OT gene list with top listed 20 genes with best global score (global OT score >0.55). The second model group, named “*PWAS-genetics”* was composed of the genes with OTG which ranked genes only through evidence from genetic association (GA). We analyzed 18 such genes with a GA score >0.6). The dominant and recessive PWAS scores for each of these genes were calculated for each individual in the UKB to create the functional-based PWAS features. Surprisingly, the PWAS features that are based on gene-relevance knowledge from OT, did not improve the AUC across all models (Supplementary **Table S3)**. Using only the PWAS data as features still carries substantial predictive power (AUC ∼0.69) which is comparable to using *“only_genetic”* features (AUC ∼0.7). Based on the results, we concluded that from genetic perspective, it is mostly the APOE features that boost the model quality across all training results, and these genes and allele are already incorporated in any of the genetics used (GWAS, PWAS or both).

### 3.5. Feature importance and interpretability

We analyzed the SHAP feature importance from *“all”* model across 10 iterations (Supplementary **Table S4).** The most significant feature identified was the rs429358, a known variant of APOE. This was followed by *“Filtered_ICD10*”, *“Age_last_episode_depression”*, *“Age_stop_smoking”* and “*Medication”*, which are relatively stable across the trained models (see standard deviation (Std) in Supplementary **Table S4**. As expected, the less important features are also generally less stable. Note that some of the top listed features are signified by strong dependency among them (e.g., “*Meds_cholesterol_hypertension_diabetes*” and *“Medication”*). We further evaluated the contribution of each feature on a selected iteration of the *“all”* model using an explainable AI tool of SHAP.

**Fig. 3A** shows the top 20 features ranked by SHAP. We analyzed the genetic feature importance across various models trained on the dataset. The heatmap (**Fig. 3B**) visually presents the importance of different features for each model, providing a comprehensive overview of how each feature contributes to the predictive power of the models. The strongest feature is the rs429358, followed by the rs7412, both are APOE variants. Interestingly, the rs429358 is more important for female models (mean SHAP importance >0.5) while for males, the value SHAP value is <0.4. SNP rs117618017 demonstrates only a moderate importance across different models, with a peak in the*“only_genetics”* (0.029) and the *“selected_icd10_all”* (0.017) models.

**Figure 3.**
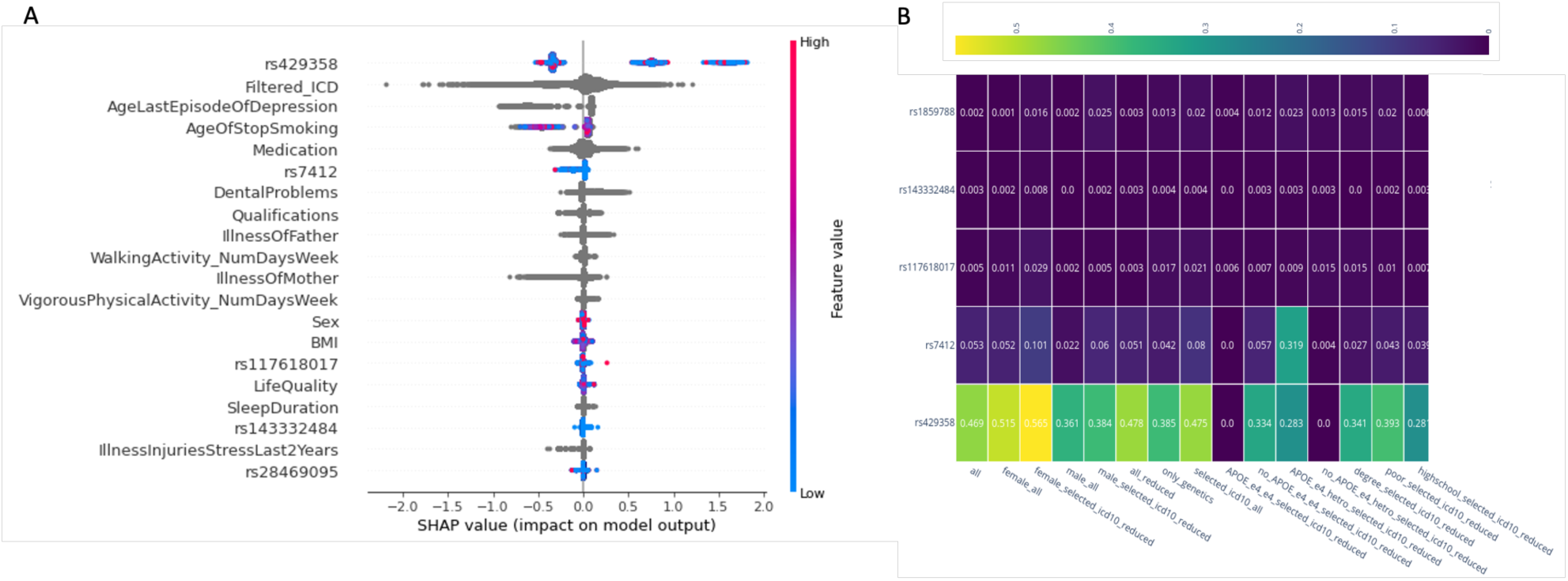
Feature importance of AD predictive risk model. **(A)** Top 20 features from the “all” model selected iteration using SHAP. The most informative feature is at the top. Each dot in the plot represents a subject patient’s feature value for that variable (vertical axis). The values reported show the contribution of each of the feature to the model outcome (i.e., AD), where the color reflects the scale of the feature’s value. Color shows whether that variable is high (red) or low (blue) for that observation. Gray depicts no data or categorical, textual features. Note that the genetic features (e.g., rs7429358, rs7412) can only take discrete values (0, 1 and 2). **(B)** Heatmap of mean SHAP importance of the 5 most influential genetic features used in models training. Importance values are rounded with 2-digit precision.

The importance of the others SNPs, such as rs143332484 (**Fig. 3B**) and rs1859788 is lower but still noteworthy importance in some models. The dominating trend is that other SNPs, including rs769452 and rs146723120, show minimal importance across most models. Overall, the heatmap highlighting the significant role of certain SNPs, particularly rs429358 and rs7412, in influencing model outcomes. We concluded that APOE variants are of utmost importance for the AD prediction outcome but the contribution of other genetic variants is negligible in improving any of the predictive models. The results emphasize the need for further investigation into these SNPs to better understand their biological implications and their potential use in predictive modeling.

### 3.6. Health records boost the AD model and exposes sex differences

The analysis of ICD-10 feature importance across various models reveals insights into the predictive power of the diagnostic codes. The heatmap in **Fig. 4A** shows that the most significant feature identified across several models is E11.9 (Diabetes without complications). Another notable feature is I10 Essential (primary) hypertension, which shows substantial importance across various models, especially in the *“selected_icd10_all”.* Note that ICD-10 I10 provided additional values for models of both sexes. The R07.4 (Chest pain, unspecified), I20.9 (Angina pectoris, unspecified), and E78.0 Pure hypercholesterolemia carry considerable importance in only some but not all models. Overall, the heatmap illustrates the variability in ICD-10 feature importance across different models, with some features consistently showing high importance, while the contribution of other diseases is negligible. These findings emphasize the need for a combined and weighted set of features to enhance model accuracy and provide valuable insights into the health determinants of AD within the tested population (for extensive analysis of models and ICD-10 importance, see Supplementary **Fig. S2**).

**Figure 4.**
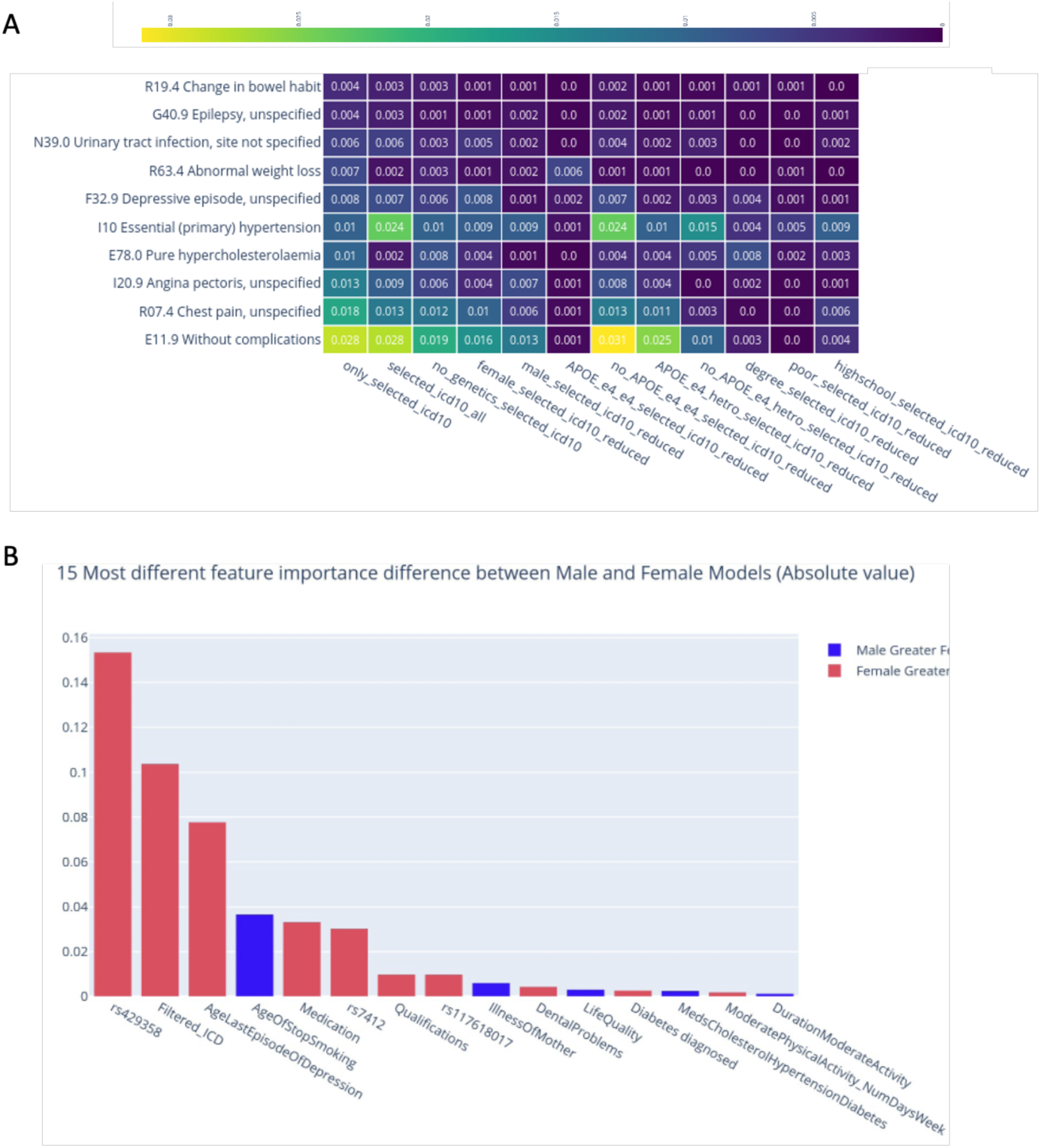
Interpretation of AD predictive risk model by the feature properties. **(A)** Heatmap of mean SHAP importances of the selected ICD-10 features used in model training. ICD-10 features are sorted by the only_selected_icd10 model mean feature importance. Importance values are rounded with 2 digit precision. (**B)** Histogram of the most different features between “male_all” and “female_all” models. The values of the most different feature importance are shown. Colored in red and blue are features that are more important in the female and male models, respectively.

We compared the mean feature importance of the *“male_all”* and “*female_all”* models to emphasize instances that maximize the different features that carry sex-dependent differences (**Fig. 4B**). Interestingly, the rs429358 APOE variant is relatively more important for the female model than the male model, followed by the filtered ICD-10 diagnosis, the age of last episode of depression, medication, and rs7412 APOE variant. Age of stop smoking has a greater mean SHAP importance value for males. The tail of the features has a difference of less than 0.01mean SHAP importance and will not be further discussed.

### 3.7. Models for vascular dementia (VaD) outperform AD and dementia models

AD is usually diagnosed through MRI and brain imaging with the majority of dementia cases are unspecified, and may occur as a mixed dementia [49]. **Fig. 5A** shows the different dementia by ICD-10 indexing. The dementia models consist of any patients that have at least one positive diagnosis for the ICD-10 codes of F00, F01, F02 or F03. Together there are 6,227 positive samples (the sum of 2,216, 33, 275, 411, 807 and 2,485 samples). The Non-vascular dementia models consist of patients with F00, F02 or F03 with no F01 diagnosis (2,485 + 2,216 = 4,701 positive samples). Vascular dementia (VaD) models consist of patients with F01 (807 + 33 + 275 + 33 = 1,526 positive samples) and the Unique VaD models consist of patients with just F01 and no F00, F02 or F03 diagnosis (807 +33 = 840 positive samples).

**Figure 5.**
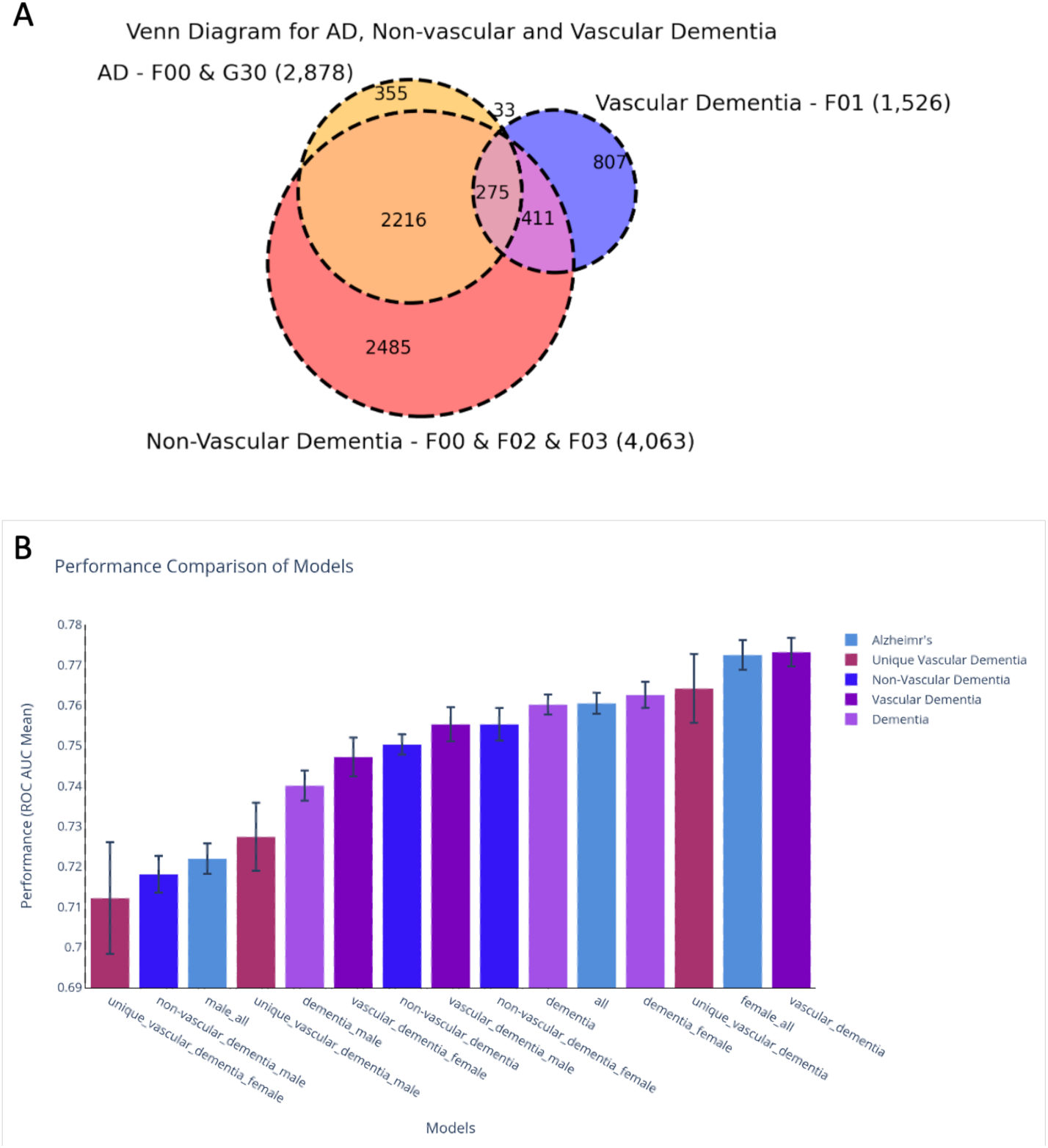
Partition of dementia cohort in UKB to subgroups by clinical ICD-10. **(A)** Venn diagram of different Dementia diagnoses. The AD model consists of patients with F00 or G30 diagnosis (355 + 2,216 + 33 + 274 = 2,878 positive samples). **(B)** Comparison of models’ performance by ROC-AUC mean of 10 different training iterations. Error bars present the standard errors. Bars are colored by their categories: AD, unique vascular dementia, non-vascular dementia, vascular dementia (VaD) and dementia.

For the goal of improving prediction by dementia subtyping, we performed model training for different diagnoses and created models for male, female and both sexes (i.e., *“all”).* The results are summarized in **Fig. 5B**. The model with the best ROC-AUC metric is for VaD. The female models are stronger than the male-centric models for AD, non-vascular dementia and dementia in general. But the male models perform better for VaD and for Unique VaD. Notably, the unique VaD model has a relatively high ROC-AUC score (0.764), but this measure is lowered for the male and female models. We suspected that the difference is due to the relatively low number of positive samples for these models (<500 samples) and relatively high standard errors. Testing the most likely explanation for the sex difference failed to support sex dependency. The limited cohort sizes dominated the results as shown by down-sampling and bootstrapping protocols. We conclude that there is no statistically significant difference between the means of the two groups and we claim that there is no sufficient evidence to suggest that the different samples come from different populations. The performance of the different models is compiled in Supplementary **Table S5**.

### 3.8. Difference in feature importance for VaD, AD, and dementia models

The features that are more important in AD models includes rs7412 APOE variant, illness_of_father, qualifications, meds_cholesterol_hypertension_diabetes and diabetes diagnosed. The features that are more important in dementia models includes BMI, life_quality (datafield 26417), duration_moderate_activity walking_activity (num days in week), rs117618017, sex, and smoking_packs_years. This list strongly proposed the importance of life style, physical activity and family history.

We observed that the unique VaD models have some features that are substantially more important relative to the other models. These includes LDL_cholesterol (average rank of 5.3 vs 34.35 and 41.05 in AD and dementia models, respectively), cardiovascular_diagnosis (average rank of 14.40 vs 43.45 and 24.50 in AD and dementia models) and seen_shrink_for nerves, anxiety, tension, depression (average rank of 21.10 vs 37.15 and 35.00 in AD and dementia models). Features that are substantially less important in unique VaD models includes Medication (average rank of 31.75 vs 5.40 and 3.60 in AD and dementia models, respectively) and dental_problems (average rank of 31.55 vs 11.80 and 11.80 in AD and dementia models, respectively). Note that APOE rs429358 is ranked first or second in all models (with Std error 0).

## 4. Discussion

AD is a progressive neurodegenerative disorder that causes memory loss and cognitive decline, accounting for ∼70% of dementia cases. Its hallmarks include the amyloid plaques and tau tangles which are typically confirmed in postmortem brains. In living patients, diagnosis relies on imaging methods that reveal brain atrophy, reduced hippocampal volume, and signs of neuronal loss [50]. Dementia itself is a broader term for conditions marked by cognitive deterioration. Fewer than half of AD patients are marked as AD isolated, most patients are diagnosed with mixed pathologies, underscoring the diagnostic complexity [49]. Currently, no reliable blood or routine lab tests exist to differentiate dementia subtypes.

This study presents a statistical machine learning-based framework for predicting AD and dementia subtypes. Using UKB clinical data, we address key methodological challenges, particularly the lack of detailed follow-up longitudinal information. Our models leverage isolated aspects of lifestyle and experiential data from thousands of datafields, without including imaging or CSF-based measures, which are expect to enhance prediction [51, 52]. Integrating multimodal data such as imaging, proteomics, and genetics [28, 53, 54], along with detailed longitudinal information [55] were confirmed to improved AD prediction. By focusing on EHR-derived features, our approach aims to support real-world healthcare systems lacking sophisticated brain imaging data. Overall, the robustness and reproducibility of our models are essential for any future clinical application. Our predictive models revealed sex-specific contributing variables, offering insights that may inform personalized medical approaches. A key strength of this study is the detailed analysis of feature importance for the performance of alternative models. Feature importance correlations across AD, VaD, and non-vascular dementia ranged from 0.64 (AD with unique VaD) to 0.75 (AD and dementia; Supplementary **Table S5**). The VaD predictive model outperformed all others, including the "female_all" model (**Fig. 5B**). Despite the overlap in clinical manifestations of AD and VaD, the cause of VaD is reduced blood flow to the brain due to strokes or other conditions that damage blood vessels. This form of dementia can develop suddenly (e.g., following a stroke) or gradually from existing chronic vascular disease. While neurodegeneration is a progressive and long-lasting condition, in VaD, symptoms may worsen abruptly. We showed that the key difference in the cause of AD versus VaD is reflected in the analysis of the main features of each model.

An important aspect of our research is the utility of the age-matching protocol. Although we used the age of diagnosis as an alignment metric, this definition is somewhat arbitrary, as AD diagnoses are often influenced by health insurance policy, available support, and other factors rather than strict clinical determinants. Most models identify age as the primary factor, followed by sex and ethnicity (e.g., [55]). Instead, we explicitly used sex as separate groups for modeling to fully address the partition of key factors by sex (Fig. 4B). To minimize statistical bias and ensure robust interpretations, we routinely applied down-sampling techniques paired with bootstrapping to avoid any misinterpretation due to potentially small sample sizes and imbalanced groups. We further leveraged AI technology to model complex interactions, which are often nonlinear and challenging to test directly due to the rapid growth of possible combinations.

We found that the strongest (and almost exclusive) genetic contribution to the models is based on the combination of the APOE alleles. The dominant effect of the two APOE alleles is consistent with current knowledge, where the ε4 allele (i.e., rs429358_C, rs7412_C) is associated with a significantly increased risk of AD. The small number of individuals with two ε4 alleles (homozygous) can have an 8-to-12 times increased risk compared to people without ε4. In contrast, the ε2 allele (i.e., rs429358_T, rs7412_T) appears to be protective. These two extreme combinations are captured by our models. The contribution of a few additional SNPs that were selected was negligible [56]. An unresolved issue concerns the limited contribution of results from genetic association studies (excluding APOE). It is worth noting that no consensus is established regarding specific genes or variants associated with AD and dementia. The models that included pre-calculated polygenic risk score (PRS) from UKB showed negligible contribution to the genetic models. Nevertheless, a benchmark for selecting optimal PRS for modeling AD emphasized the sensitivity of careful methodological choices in genetic risk stratification [57]. While only unequivocally APOE gene was associated with AD risk, recent approach for AD genetics based on rare variants from whole exomes and whole genomes were not included in this study.

There are numerous limitations in this study that need to be addressed. Primarily, the relatively low prevalence of AD patients suggests a selection bias during recruitment. The underrepresentation of diseases such as cancer was confirmed in the UKB relative to the general population [58]. To account for such bias, further validation with independent datasets is necessary. Testing alternative biobanks (e.g., All of Us) will be pursued in the future to assess the success of our model’s transferability. Another obvious limitation concerns the cohort used. We trained the model on UKB data from 2019, and in 2023 there were additional ∼1,500 patients diagnosed with AD that were diagnosed in this time window from the recruitment date. We therefore anticipate relatively large amounts of false negatives that potentially reduced the quality of our models. Our findings underscore the significant role of education in lowering the risk of AD, likely through its association with healthier lifestyles and improved health management. Higher educational attainment has been linked to delayed onset of symptoms and protective changes in brain structure [59]. However, this protective effect was notably reduced in VaD, particularly in cases classified as uniquely VaD, suggesting distinct underlying etiologies. For example, strong lipidomic signatures observed in VaD were absent in AD, reinforcing the specificity of certain risk factors.

### CRediT authorship contribution statement

A.S. performed the analysis, developed the predictive models, created the visualization and the supplementary materials. M.L. wrote the initial draft. M.L. contributed in conceptualization, mentoring and coordination. A.S and M.L. contributed to final writing, editing and finalizing the manuscript.

## Funding

This study was supported in part by the National Alopecia Areata Foundation (NAAF) on Genetics of alopecia areata (M.L.) and the Ministry of Science and Technology (MOST) Heart-Kineret (2024). A student fellowship by the Center for Interdisciplinary Data Science Research (CIDR, 3035000440).

### Ethics

The study was approved by the University Committee for the Use of Human Subjects in Research Approval number 12072022 (July 2025). This study uses the UK-Biobank (UKB) application ID 26664 (Linial lab).

## Data Availability

This study uses the UK-Biobank (UKB) application ID 26664 (Linial lab).

## Acknowledgments

We thank the member of the Linial’s lab for their support throughout this work. Special thanks to Roei Zucker that helped with UKB data management and technical support. Special thanks to Shachar Arzi (Dept of Cognitive and Brain Sciences, The Hebrew University) and his team for introducing an expert view of AD and dementia.

## Supplementary Materials

Supplementary Text S1-S3.

Supplementary Tables S1-S5.

Supplementary Figures S1-S2.

### Supplementary Text S1

#### Hyperparameter configuration

CatBoost models were trained with the following key hyperparameters: (i) Maximum tree depth: 6; (ii) Maximum iterations: 1,000; (iii) Early stopping rounds: 15 (training terminated if validation AUC showed no improvement for 15 consecutive iterations); (iv) Evaluation metric: area under the ROC curve (ROC-AUC). (v) Class balancing: positive class weighted by the ratio of negative to positive samples; (vi) Learning rate: default (automatically determined by CatBoost); (vii) Categorical feature handling: Native CatBoost encoding; (viii) Random seed: varied across iterations (0-9) for robustness assessment.

#### Supplementary Text S2

##### Algorithm 1 Sample Ages Distribution Training Set

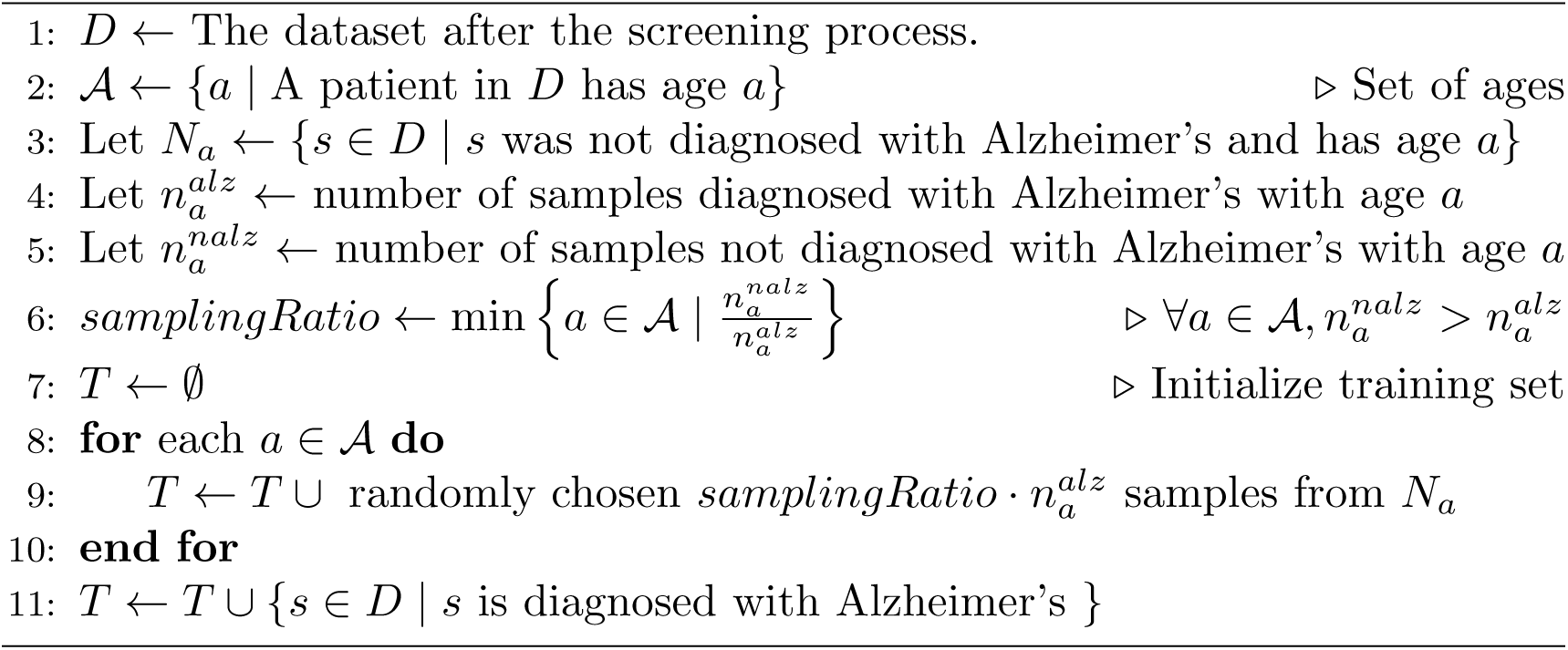

### Supplementary Text S3

#### PWAS gene-based functionality score

PWAS attempts to capture genetic variants with inference for the impact of any variant that within a occurs coding region. The underlying notion is that accumulating the score of proteins that were severely damaged allows to statistically assess the significant of any affected genes on the phenotype in a case-control setting [41]. The PWAS scores are based on quantifying the impact inference of all reported protein alterations. PWAS score are based on FIRM, a pre-trained machine-learning model, is used to estimate the extent of the damage caused to each protein in the entire proteome for each person, irrespective to the studies phenotype [40].

PWAS considers missense, nonsense, frameshift, in-frame indel, and canonical splice-site variants. It intuitively reflects the likelihood that the affected gene will retain its function in the presence of the variant. PWAS creates a protein function damage matrix that addresses all coding genes in the human proteome (k genes), as a result of the observed variants (m), for each of the n individuals (typically k < < m). The per-variant damage predictions are then aggregated at the gene level to create two distinct scores for each participant’s gene for a dominant and recessive inheritance model. The compilation of the PWAS results for >520 phenotypes, including AD is available in PWAS-Hub portal [42].

In this study we considered OTG as a most robust gene candidates per phenotype (based on the compilation from the GWAS catalog). For each candidate gene, we provided the FIRM score that were used for the AI model as quantitative feature. FIRM scores range between 0 and 1, where a score of 0 for a specific person’s gene represents damaged gene with a predicted loss of all functionality, and a value of 1 represents full functionality. Note that PWAS provides two inheritance modes for dominant or recessive effects per each phenotype.

**Table S1:** Statistical test of age distributions differences between AD and controls before and after the age-maching protocol

| **AD vs. Control (original)** |  |  |
| --- | --- | --- |
| **Test** | **statistic** | **p-value** |
| KS-test | 0.481 | 0 |
| Mann–Whitney U | 1078213763 | 0 |
| T-test | 53.297 | 0 |

| **AD vs. Control (age-matched)** |  |  |
| --- | --- | --- |
| **Test** | **statistic** | **p-value** |
| KS-test | 9.05E-05 | 1 |
| Mann–Whitney U | 104106113.5 | 0.992 |
| T-test | -0.017 | 0.985 |

**Table S2:** Top 15 significant ICD10 (FDR <0.05) from Fisher’s Exact test of AD and control groups.

| **ICD-10 Diagnosis** | **ALZ freq** | **control freq** | **odds ratio** | **Fischer’s p-value** | **FDR** | **z-value** |
| --- | --- | --- | --- | --- | --- | --- |
| I10 Essential (primary) hypertension | 0.29 | 0.15 | 2.44 | 1.25E-90 | 4.10E-87 | 0.58 |
| E78.0 Pure hypercholesterolaemia | 0.15 | 0.06 | 2.61 | 7.00E-59 | 1.15E-55 | 0.69 |
| I20.9 Angina pectoris, unspecified | 0.09 | 0.03 | 3.31 | 2.96E-57 | 3.25E-54 | 1.13 |
| E11.9 Without complications | 0.10 | 0.03 | 3.10 | 2.54E-54 | 2.09E-51 | 0.99 |
| I25.1 Atherosclerotic heart disease | 0.09 | 0.04 | 2.68 | 3.76E-40 | 2.48E-37 | 0.74 |
| I25.9 Chronic ischaemic heart disease, unspecified | 0.06 | 0.02 | 3.47 | 3.45E-39 | 1.89E-36 | 1.23 |
| H26.9 Cataract, unspecified | 0.06 | 0.02 | 2.91 | 1.04E-33 | 4.90E-31 | 0.88 |
| Z86.4 Personal history of psychoactive substance abuse | 0.09 | 0.04 | 2.36 | 2.11E-31 | 8.68E-29 | 0.53 |
| Z37.0 Single live birth | 0.00 | 0.02 | 0.00 | 3.11E-29 | 1.14E-26 | -0.94 |
| R07.4 Chest pain, unspecified | 0.10 | 0.05 | 2.18 | 4.07E-29 | 1.34E-26 | 0.42 |
| I48 Atrial fibrillation and flutter | 0.06 | 0.02 | 2.64 | 2.58E-27 | 7.72E-25 | 0.71 |
| Z92.2 Personal history of long-term (current) use of other medicaments | 0.06 | 0.02 | 2.54 | 6.91E-24 | 1.89E-21 | 0.65 |
| Z86.7 Personal history of diseases of the circulatory system | 0.06 | 0.03 | 2.38 | 9.03E-23 | 2.29E-20 | 0.54 |
| K57.3 Diverticular disease of large intestine without perforation or abscess | 0.08 | 0.04 | 2.10 | 1.28E-21 | 3.00E-19 | 0.37 |
| I25.8 Other forms of chronic ischaemic heart disease | 0.04 | 0.02 | 2.73 | 1.31E-20 | 2.87E-18 | 0.76 |

**Table S3.** Model performance summary with OT genes and PWAS scoring.

| **Model (Abbreviated name)** | **AUC (mean)** | **AUC (std)** |
| --- | --- | --- |
| female_all | 0.773 | 0.012 |
| female_all_PWAS_OT-genetics | 0.769 | 0.014 |
| female_all_PWAS_OT-global | 0.767 | 0.013 |
| all | 0.761 | 0.008 |
| all_PWAS_OT-global | 0.760 | 0.008 |
| all_PWAS_OT-genetics | 0.757 | 0.008 |
| male_all_PWAS_OT-genetics | 0.723 | 0.015 |
| male_all_PWAS_OT-global | 0.722 | 0.018 |
| male_all | 0.722 | 0.012 |
| only_PWAS_OT-genetics | 0.693 | 0.009 |
| only_PWAS_OT-global | 0.692 | 0.009 |

**Table S4.**
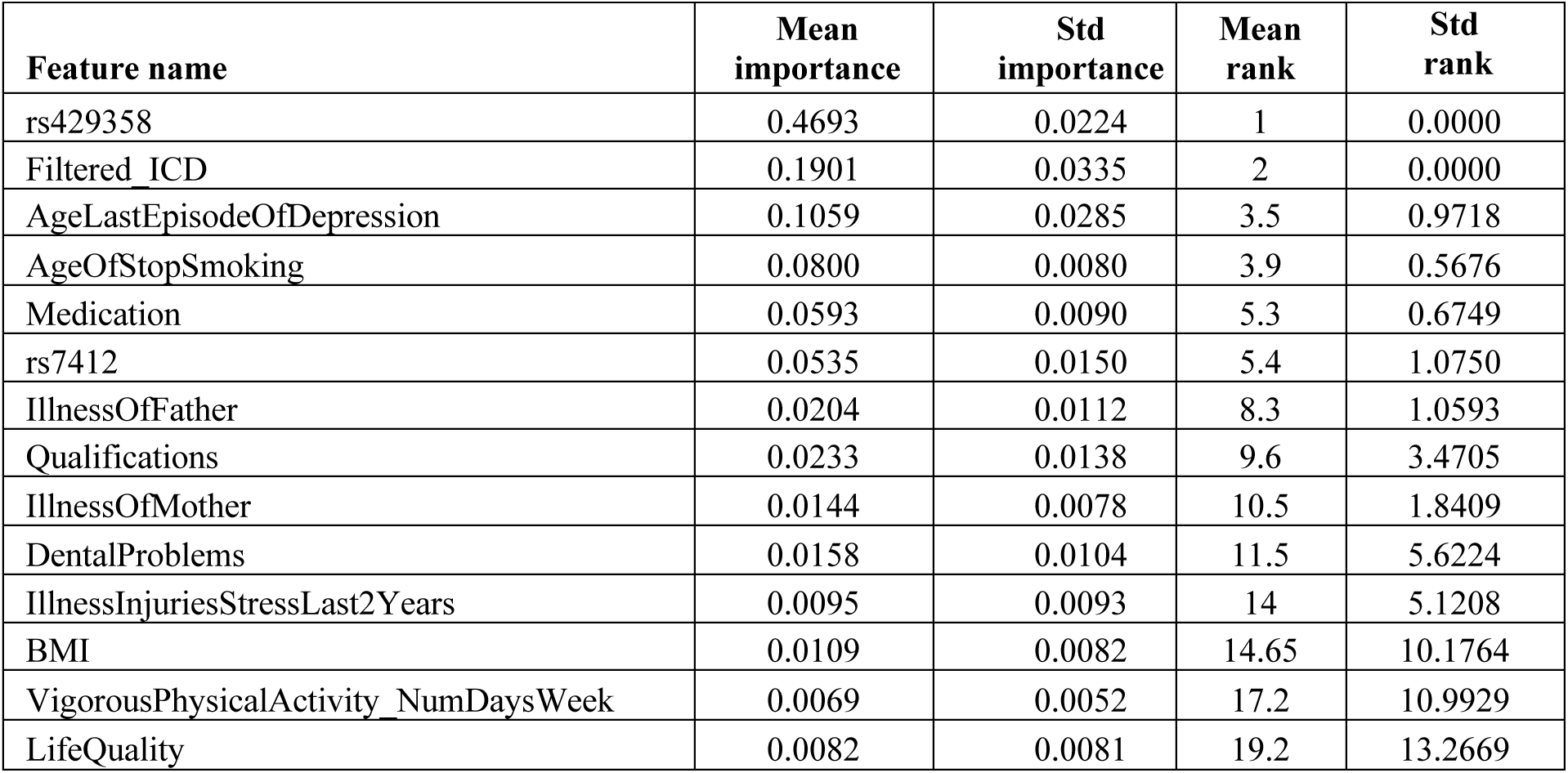
Feature importance for the top features of the “all” model summary results of 10 iterations.

**Table S5:** Models metrics summary of AD and Dementia models.

| model | AUC mean | Accuracy mean | Precision mean | Recall mean | F1 mean | # of samples | # of positive |
| --- | --- | --- | --- | --- | --- | --- | --- |
| vascular_dementia | 0.773 | 0.764 | 0.519 | 0.701 | 0.476 | 88924 | 1526 |
| Alzheimer's female | 0.773 | 0.758 | 0.540 | 0.710 | 0.514 | 40343 | 1475 |
| unique_vascular_dementia | 0.764 | 0.771 | 0.510 | 0.690 | 0.459 | 90058 | 840 |
| dementia_female | 0.763 | 0.730 | 0.561 | 0.697 | 0.543 | 42884 | 2948 |
| Alzheimer's all | 0.761 | 0.751 | 0.539 | 0.694 | 0.512 | 75232 | 2878 |
| dementia | 0.760 | 0.723 | 0.564 | 0.693 | 0.546 | 82814 | 6227 |
| vascular_dementia_male | 0.755 | 0.759 | 0.520 | 0.686 | 0.477 | 44357 | 893 |
| non-vascular_dementia_female | 0.755 | 0.742 | 0.551 | 0.690 | 0.531 | 41227 | 2315 |
| non-vascular_dementia | 0.750 | 0.729 | 0.551 | 0.684 | 0.527 | 78339 | 4701 |
| vascular_dementia_female | 0.747 | 0.715 | 0.513 | 0.685 | 0.448 | 43496 | 633 |
| dementia_male | 0.740 | 0.709 | 0.561 | 0.676 | 0.540 | 40252 | 3279 |
| unique_vascular_dementia_male | 0.727 | 0.765 | 0.509 | 0.655 | 0.456 | 48135 | 482 |
| Alzheimer's male | 0.722 | 0.747 | 0.534 | 0.664 | 0.505 | 35136 | 1403 |
| non-vascular_dementia_male | 0.718 | 0.731 | 0.548 | 0.659 | 0.527 | 37401 | 2386 |
| unique_vascular_dementia_female | 0.712 | 0.725 | 0.507 | 0.661 | 0.437 | 41443 | 358 |

**Figure S1.**
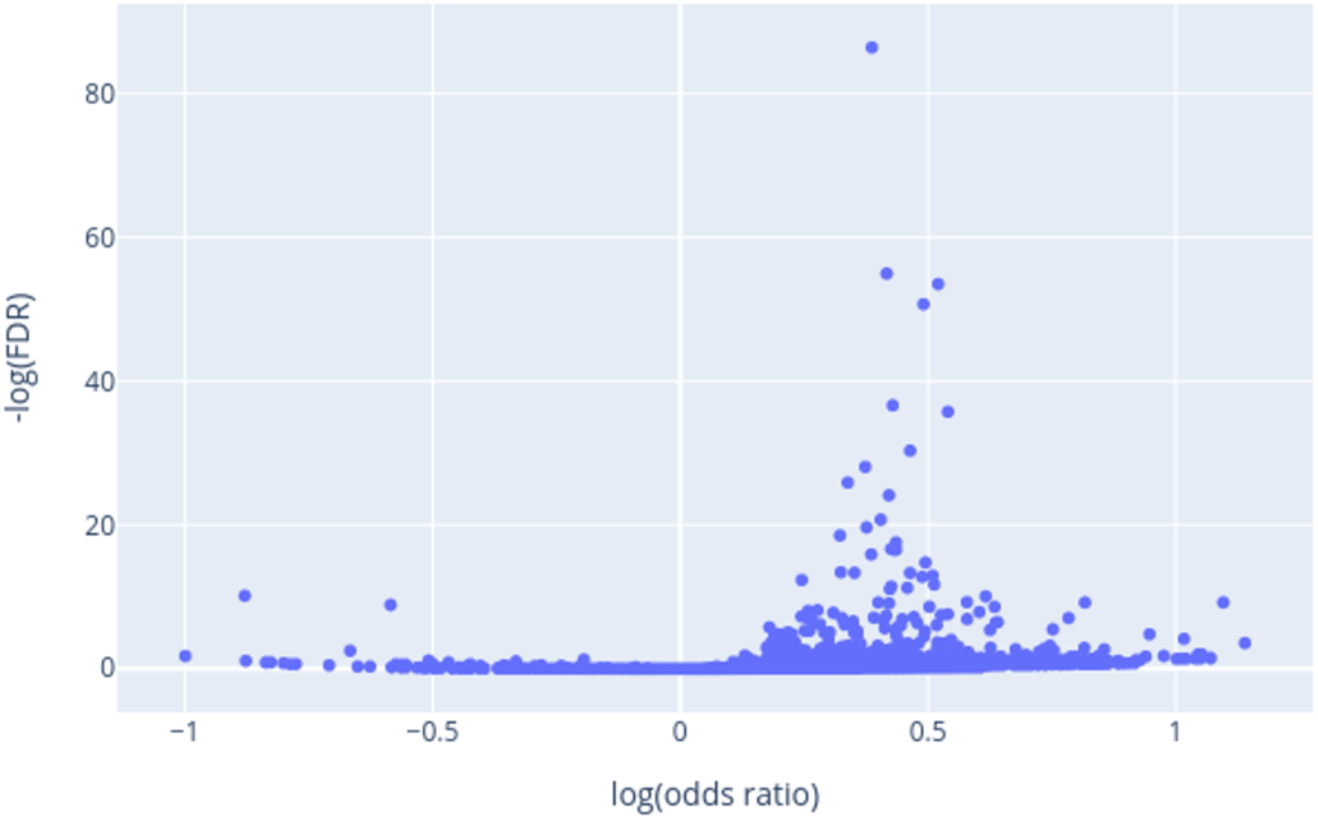
Plot of -log10(q-value FDR) vs. log2(odds ratio; OR) of Fisher’s exact test for ICD-10 diagnosis of AD and control groups. The figure captures the effect size and statistical significance.

**Fig. S2.**
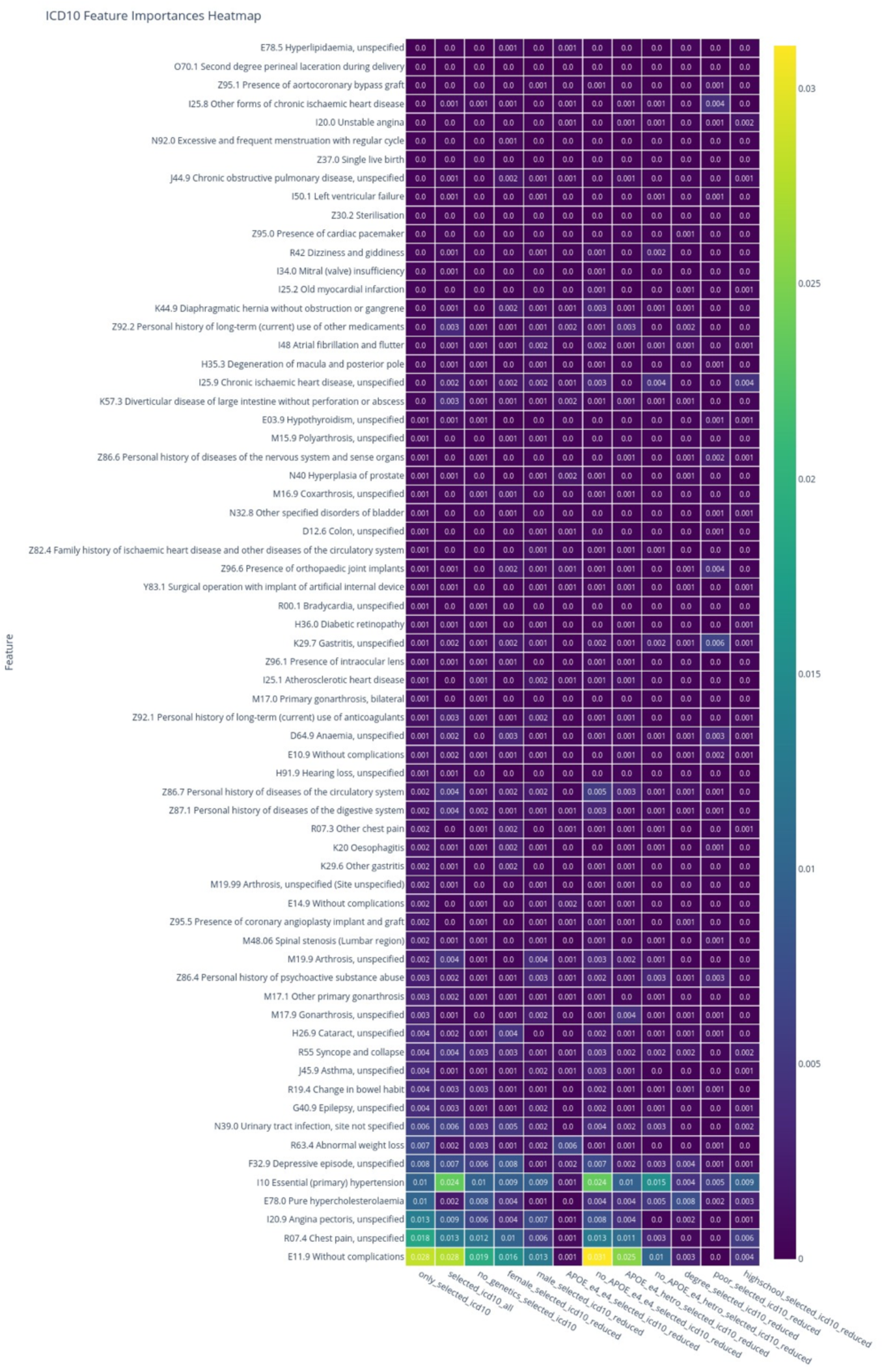
Heatmap of mean SHAP importances of all 66 ICD-10 features used in model training. ICD-10 features are sorted by the only_selected_icd10 model mean feature importance. Importance values are rounded with 2 digit precision.

